# Trends in serotype distribution and disease severity in adults hospitalised with *Streptococcus pneumoniae* infection in Bristol and Bath: a retrospective cohort study, 2006-2022

**DOI:** 10.1101/2023.03.30.23287917

**Authors:** Catherine Hyams, Robert Challen, David Hettle, Zahin Amin-Chowdhury, Charli Grimes, Gabriella Ruffino, Rauri Conway, Robyn Heath, Paul North, Adam Malin, Nick A Maskell, Philip Williams, O. Martin Williams, Shamez N Ladhani, Leon Danon, Adam Finn

**Affiliations:** Academic Respiratory Unit, Learning and Research Building, Southmead Hospital, Bristol, UK; Bristol Vaccine Centre, University of Bristol, Bristol, UK; Engineering Mathematics, University of Bristol, Bristol, UK; Department of Microbiology, Bristol Royal Infirmary, Upper Maudlin Street, Bristol, UK; Immunisation Department, UK Health Security Agency (UKHSA), London, UK; Department of Respiratory Medicine, The Royal United Hospital, Bath, UK

**Keywords:** Streptococcus pneumoniae, pneumococcus, serotypes, bacterial infection, respiratory infection

## Abstract

Ongoing surveillance is essential to inform policy decisions and monitor serotype replacement following pneumococcal conjugate vaccination (PCV) deployment. We report serotype and disease severity trends in this retrospective cohort of hospitalised adults in Bristol-Bath, 2006-22. Of 1686 invasive pneumococcal disease (IPD) cases, 1501 (89.0%) had known serotype. We also identified 2033/3719 cases of non-IPD. IPD declined sharply during the early COVID-19 pandemic. Over 2022 it gradually returned to pre-pandemic levels. Disease severity also changed throughout this period: CURB65 severity and inpatient mortality decreased whilst ICU admissions increased. PCV7 and PCV13-serotype IPD decreased from 2006-09 to 2021-22. However, significant residual PCV13-serotype IPD remains, representing 21.7% [15.5-29.6] of 2021-22 cases, highlighting that significant adult PCV-serotype disease still occurs despite 17-years of paediatric PCV usage in the UK. We found increased proportions of serotype 3 and 8 IPD, whilst 19F and 19A re-emerged. In 2020-22, 68.2% IPD cases were potentially covered by PCV20.

**Article Summary:** We observed significant serotype shifts but perseverance and re-emergence of some serotypes covered by PCVs over this 17-year retrospective study, which found considerable adult pneumococcal disease attributable to PCV-serotypes despite high uptake of paediatric PCV.

## INTRODUCTION

*Streptococcus pneumoniae* remains the leading bacterial cause of community-acquired pneumonia (CAP), despite widespread use of effective pneumococcal vaccines with over 100 recognised pneumococcal serotypes. In the UK, PPV23 (unconjugated 23-valent pneumococcal polysaccharide vaccine) is offered to all adults aged ≥65 years (y), and persons aged at least 2y who are at increased risk of pneumococcal disease. In September 2006, a 7-valent pneumococcal conjugate vaccine (PCV7) was implemented into the national childhood immunisation programme and replaced with a 13-valent PCV (PCV13) in April 2010. The PCVs were given at a 2+1 schedule but replaced with a 1+1 schedule in April 2020 [1]. Because PCVs prevent carriage acquisition in addition to protection against disease, both PCVs had a large and significant direct and indirect (herd) impact on pneumococcal disease caused by the respective serotypes [2–4]. By 2016/17, PCV7 serotype disease had virtually disappeared in children and decreased significantly in adults. Invasive pneumococcal disease (IPD) due to PCV13 serotypes has also declined substantially, but subsequently plateaued, with a residual incidence of 8 IPD cases per 100,000 population in England [5–7]. At the same time, pneumococcal cases due to non-PCV13 serotypes increased across all age groups, especially in older adults, resulting in no net reduction in total IPD cases in older adults in 2016/17 compared to the pre-PCV13 period: a phenomenon known as serotype replacement [5–7]. Monitoring these effects nationally is vital for planning of healthcare utilisation and developing new preventive strategies, including use of higher-valent pneumococcal vaccines [8,9].

However, the impact of serotype replacement has not been reported in the USA; data suggests that the incidence on non-vaccine serotype disease has remained stable, in both children and older adults [10]. Understanding the reasons for differences seen in studies from the US and UK is important, since serotype replacement is a threat to the effectiveness of current vaccine programmes and PCV scheduling remains a policy decision area. It remains unclear why these differences occur; however, several factors may operate such as methodological differences in surveillance approaches, exposure to pneumococcal transmission, risk factor profiles between patient populations, and serotype interactions [11]. Notably, small differences in carriage prevalence and clonal lineages may result in significantly different rates of IPD, as serotype-specific invasiveness varies by orders of magnitude.

In England, the UK Health Security Agency conducts national IPD surveillance, which includes limited data on non-invasive pneumococcal disease, clinical phenotype and disease severity [5–7]. Evidence from a large pneumococcal pneumonia cohort in Nottingham, UK suggests there are relatively few differences between patients with PCV13 and non-PCV13 serotype respiratory infection [12]. Nevertheless, changing serotype distribution could result in changes in pneumococcal disease phenotypes, which may have implications for utilisation of available polyvalent serotype-specific pneumococcal vaccines. Furthermore, the COVID-19 pandemic has disrupted the epidemiology of multiple respiratory infections [13,14], and provided new insights into viral-bacterial-host interactions. Many countries, including the UK, implemented measures such as social distancing and school closures, intended to decrease SARS-CoV-2 transmission and alleviate pressure on healthcare services [15]. These measures reduced the transmission of other respiratory pathogens [14], but it is unclear to what extent they have disrupted pneumococcal transmission [16,17]. These measures may also have caused changes in serotype distribution of pneumococcal infection, and disease.

In this retrospective cohort study conducted at three large NHS hospitals, which represent all secondary care provision within a defined geographical area, we examined the trends in pneumococcal serotype distribution in adults following PCV7 and PCV13 implementation into the childhood immunisation programme and the effect of the COVID-19 pandemic over the first three years. We report confirmed pneumococcal disease incidence during 2006-2022, both overall and by vaccine serotypes, and assess trends in severity of pneumococcal disease in hospitalised adults.

## METHODS AND MATERIALS

### Study design

A retrospective cohort study was conducted including all patients aged ≥16 years that were admitted to any of three large UK NHS hospitals in southwest England - University Hospitals Bristol and Weston, North Bristol and The Royal United Hospital (Bath) NHS Trusts. The study covered the period from 01/01/2006 to 31/12/2022 and included patients with a confirmed microbiological diagnosis of pneumococcal infection. These hospitals provide all secondary care within a defined geographical area with 100,000 unplanned adult admissions annually, including the regional cardiothoracic, pleural, respiratory specialist, and general medical and respiratory services.

Eligible cases were identified retrospectively by searching the Laboratory Information Management System (LIMS) database (Clinisys WinPath Enterprise). *S. pneumoniae* was confirmed through culture or PCR from a sterile site at a central laboratory using standard microbiological techniques, the former confirmed with API®-20 Strep (BioMérieux, UK) or MALDI-TOF (matrix-assisted laser desorption/ionisation/time of flight) mass spectrometry (Bruker, UK). A positive pneumococcal urinary-antigen test [UAT] (BinaxNOW®, Alere, UK) was also considered confirmative of pneumococcal infection. Patients were included if they tested positive on any or all tests. Confirmed cases were linked with the UK Health Security Agency (UKHSA) national reference laboratory database to obtain serotype data, which were collected at the end of the study to avoid risk of bias in clinical data collection. Clinical records were reviewed at each hospital and data were recorded in a standardised manner, including laboratory and radiological investigations. Patient observations within 24 hours of clinical presentation with a pneumococcal infection were also recorded, and vaccination status was established using electronically-linked General Practitioner (GP) records. The CURB65 severity score on admission was calculated for each clinical episode, and clinical outcomes, including length of hospitalisation and Intensive Care Unit [ICU] admission, were recorded. Inpatient mortality (i.e patient death before discharge) was determined through review of the medical records, and used as a marker of case fatality. All adults were managed at the discretion of the admitting clinical team.

### Case definitions

Total pneumococcal disease included all positive cases whether identified by sterile site culture/PCR or positive UAT with clinical confirmation of sterile site infection (i.e. invasive pneumococcal disease [IPD]), or non-invasive pneumococcal disease (positive UAT only). Total cases where the serotype was identified are referred to as serotype-known disease: pneumococcal serotypes were further grouped by vaccine-serotypes: PCV7, PCV13-7, PCV15-13, PCV20-15, PCV20-13, and serotypes not contained in a PCV (non-PCV) (Supplementary Data 1). The primary infection site was derived from the managing clinician’s diagnosis. Respiratory infection included both consolidative infection (i.e. pneumonia following BTS/NICE guidelines [18]) and non-pneumonic lower respiratory tract infection.

### Analysis

Data are reported as medians and interquartile ranges (IQR) for continuous variables, or means and standard deviations where the distributions were confirmed to be normal using the Anderson Darling normality test. Categorical variables are presented as counts and percentages. Multinomial confidence intervals were calculated using Wilson’s score interval method for binomial proportions [19] which we tested on simulated data and found to be well calibrated for this problem. Baseline characteristics were compared using Fisher’s exact tests for categorical variables, the two-sample Kolmogorov-Smirnov for non-parametric continuous variables, Wilcoxon Rank Sum test for score variables, or the two-sided Student’s t-test for parametric continuous variables. There were minimal missing data, and only for categorical variables. When present they are included as a separate category prior to statistical testing.

Multinomial time series analysis was performed by fitting a single-hidden-layer neural network using the R package ‘nnet’ [20] with a time varying natural spline term for class probabilities, and a knot point for each class every two years. Binomial time series analysis was performed using a maximum likelihood approach using local polynomial regression using a logistic link function assuming the count of positives and negatives are a quasi-binomially distributed quantity using the R package ‘locfit’ according to the methods of Loader *et al.* [21] with a bandwidth equivalent to 2 years’ worth of data and a polynomial of degree 2.

Yearly estimates for the ≥16y population of clinical commissioning groups of NHS Bath and North East Somerset and NHS Bristol, North Somerset and South Gloucestershire regions were obtained from NHS Digital. Yearly point estimates were interpolated to daily estimates using a local polynomial regression and pneumococcal disease incidence was estimated as the rate of monthly admission counts using a local polynomial regression, assuming a quasi-poisson distribution, logarithmic link function and the methods of Loader *et al.* [21] as above. Admission rates were expressed as disease per 1000 person years using the interpolated population estimates. All analyses were performed using R version 4.2 [22].

### Ethics Approval

This study was approved by the Health Research Authority, UK (IRAS 265437).

### Funding

This work was supported by a National Institute for Health Research [NIHR Academic Clinical Fellowship (ACF-2015-25-002). The views expressed are those of the author(s) and not necessarily those of the NIHR or the Department of Health and Social Care.

## RESULTS

From 2006-2022, we identified 3,719 adults with pneumococcal disease with a median age of 66.1 years [IQR 50.2–78.9]. Of these, 1,840 (49.5%) were male, 1,621 (43.6%) had positive blood cultures, 2,379 (64.0%) were UAT positive and 15 (0.4%) were PCR-positive. Among the cases, 1,686 (45.3%) were IPD and 2,033 (54.7%) were non-invasive pneumococcal disease cases. Respiratory infection accounted for 92.3% (3436/3719) of pneumococcal disease (84.2% and 99.2% of IPD and non-invasive pneumococcal disease, respectively) (Table 1). Pneumococcal serotype was available for 1,501 (40%) cases. The demographics and clinical characteristics of patients with known-serotype pneumococcal disease were similar to those with unknown serotype infection (Supplementary Data 2).

**Table One:**
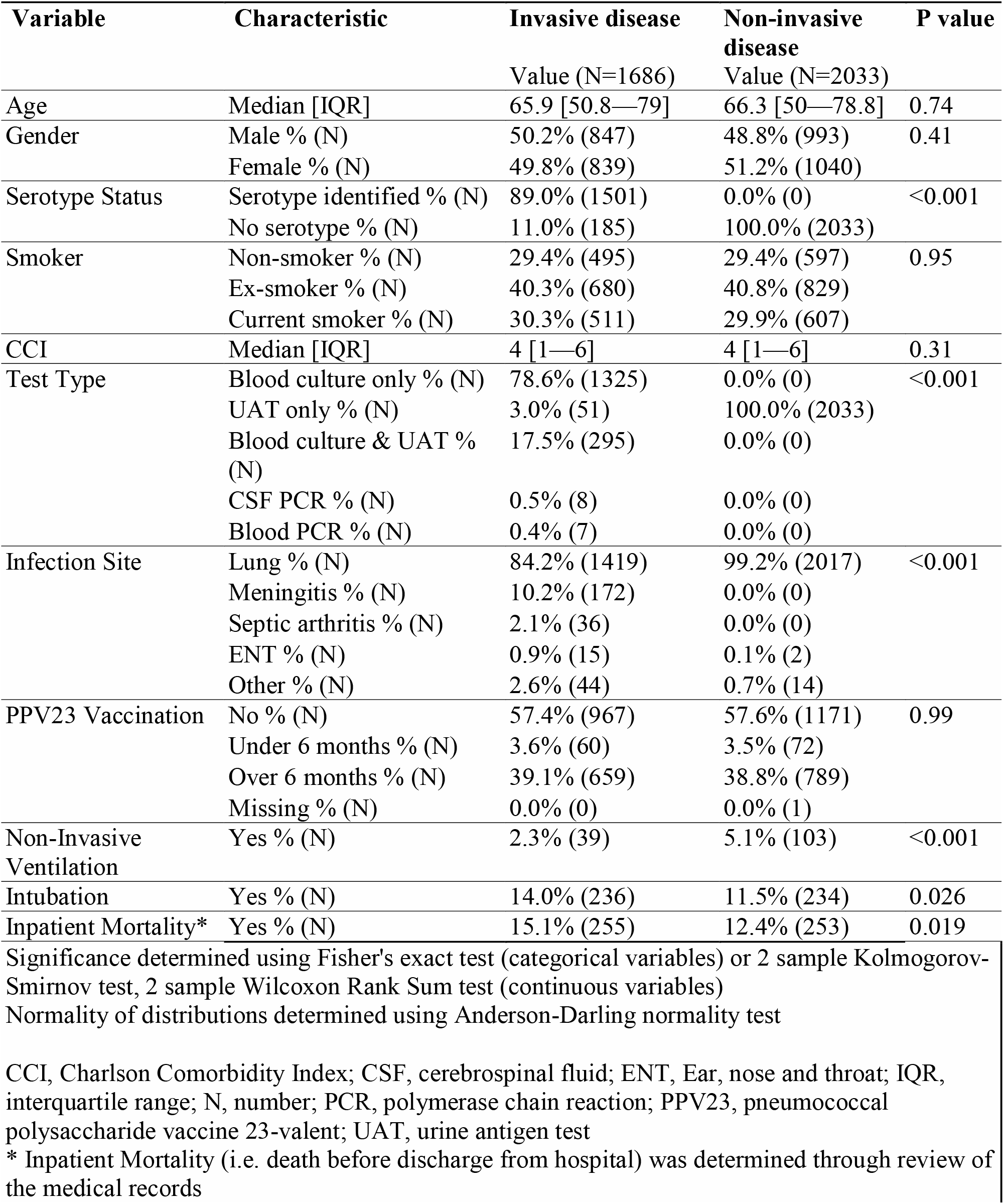
Characteristics of patients hospitalised with confirmed pneumococcal infection in Bristol, 2006-2022.

**Table Two:**
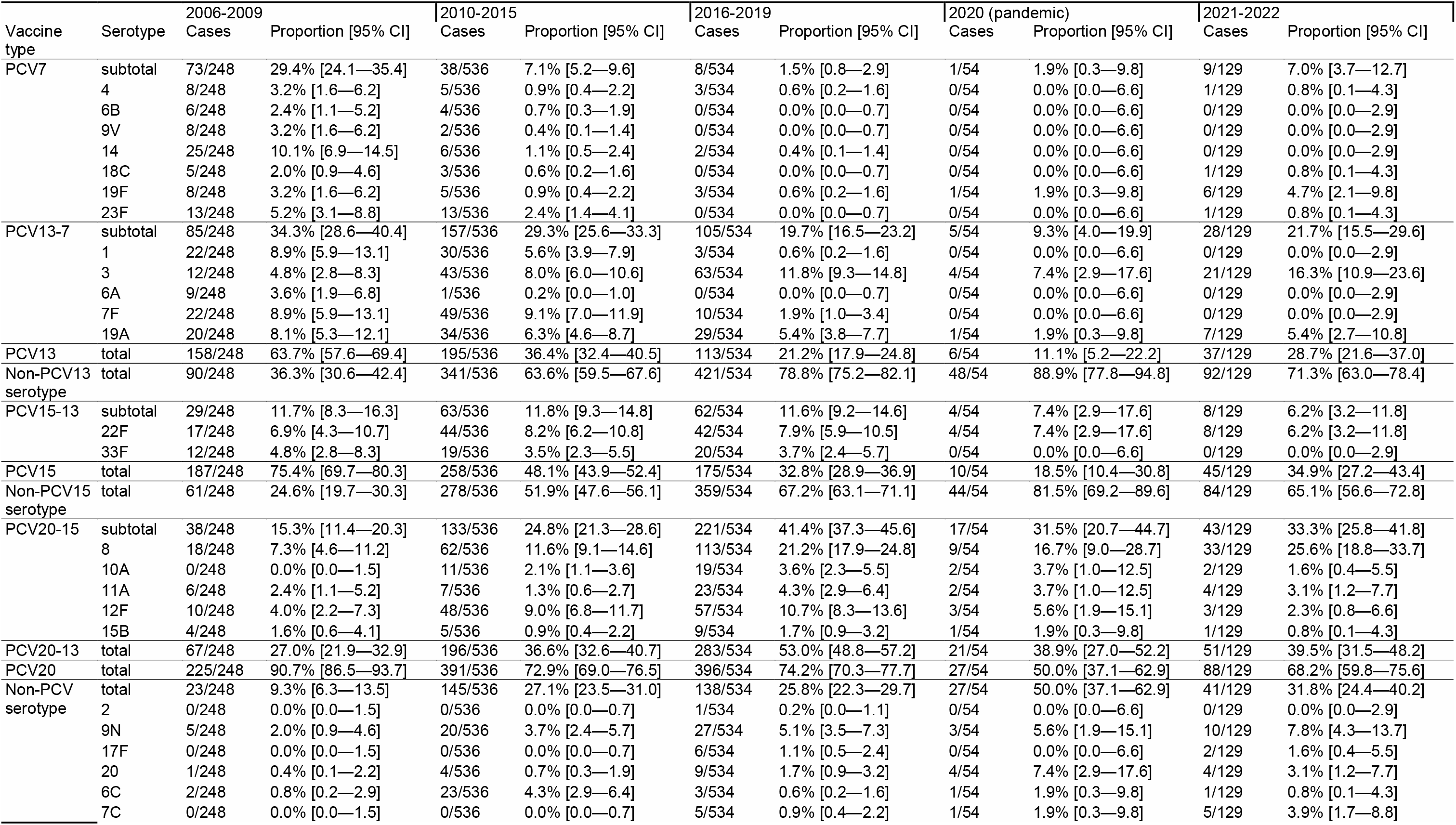
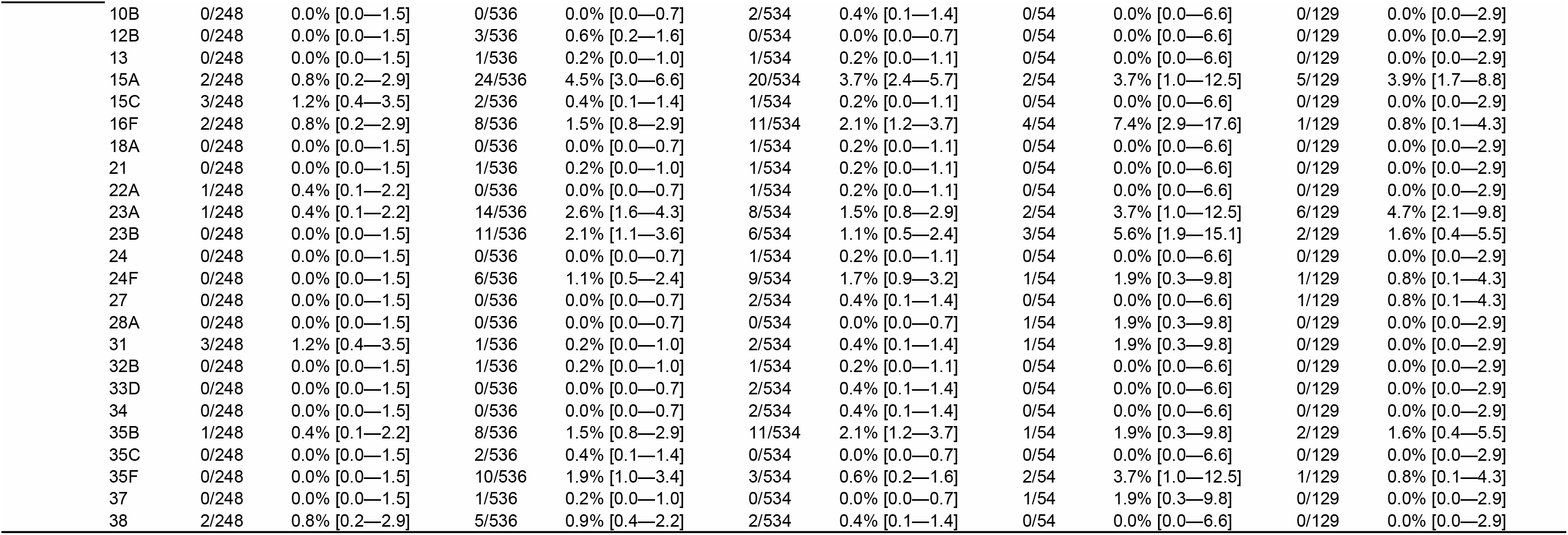
Case number and proportion of disease attributable to each pneumococcal serotype, by vaccine group, from Jan 2021 - c 2022.

| Vaccine type | Serotype | 2006-2009 |  | 2010-2015 |  | 2016-2019 |  | 2020 (pandemic) |  | 2021-2022 |  |
| --- | --- | --- | --- | --- | --- | --- | --- | --- | --- | --- | --- |
|  |  | Cases | Proportion [95% CI] | Cases | Proportion [95% CI] | Cases | Proportion [95% CI] | Cases | Proportion [95% CI] | Cases | Proportion [95% CI] |
| PCV7 | subtotal | 73/248 | 29.4% [24.1—35.4] | 38/536 | 7.1% [5.2—9.6] | 8/534 | 1.5% [0.8—2.9] | 1/54 | 1.9% [0.3—9.8] | 9/129 | 7.0% [3.7—12.7] |
|  | 4 | 8/248 | 3.2% [1.6—6.2] | 5/536 | 0.9% [0.4—2.2] | 3/534 | 0.6% [0.2—1.6] | 0/54 | 0.0% [0.0—6.6] | 1/129 | 0.8% [0.1—4.3] |
|  | 6B | 6/248 | 2.4% [1.1—5.2] | 4/536 | 0.7% [0.3—1.9] | 0/534 | 0.0% [0.0—0.7] | 0/54 | 0.0% [0.0—6.6] | 0/129 | 0.0% [0.0—2.9] |
|  | 9V | 8/248 | 3.2% [1.6—6.2] | 2/536 | 0.4% [0.1—1.4] | 0/534 | 0.0% [0.0—0.7] | 0/54 | 0.0% [0.0—6.6] | 0/129 | 0.0% [0.0—2.9] |
|  | 14 | 25/248 | 10.1% [6.9—14.5] | 6/536 | 1.1% [0.5—2.4] | 2/534 | 0.4% [0.1—1.4] | 0/54 | 0.0% [0.0—6.6] | 0/129 | 0.0% [0.0—2.9] |
|  | 18C | 5/248 | 2.0% [0.9—4.6] | 3/536 | 0.6% [0.2—1.6] | 0/534 | 0.0% [0.0—0.7] | 0/54 | 0.0% [0.0—6.6] | 1/129 | 0.8% [0.1—4.3] |
|  | 19F | 8/248 | 3.2% [1.6—6.2] | 5/536 | 0.9% [0.4—2.2] | 3/534 | 0.6% [0.2—1.6] | 1/54 | 1.9% [0.3—9.8] | 6/129 | 4.7% [2.1—9.8] |
|  | 23F | 13/248 | 5.2% [3.1—8.8] | 13/536 | 2.4% [1.4—4.1] | 0/534 | 0.0% [0.0—0.7] | 0/54 | 0.0% [0.0—6.6] | 1/129 | 0.8% [0.1—4.3] |
| PCV13-7 | subtotal | 85/248 | 34.3% [28.6—40.4] | 157/536 | 29.3% [25.6—33.3] | 105/534 | 19.7% [16.5—23.2] | 5/54 | 9.3% [4.0—19.9] | 28/129 | 21.7% [15.5—29.6] |
|  | 1 | 22/248 | 8.9% [5.9—13.1] | 30/536 | 5.6% [3.9—7.9] | 3/534 | 0.6% [0.2—1.6] | 0/54 | 0.0% [0.0—6.6] | 0/129 | 0.0% [0.0—2.9] |
|  | 3 | 12/248 | 4.8% [2.8—8.3] | 43/536 | 8.0% [6.0—10.6] | 63/534 | 11.8% [9.3—14.8] | 4/54 | 7.4% [2.9—17.6] | 21/129 | 16.3% [10.9—23.6] |
|  | 6A | 9/248 | 3.6% [1.9—6.8] | 1/536 | 0.2% [0.0—1.0] | 0/534 | 0.0% [0.0—0.7] | 0/54 | 0.0% [0.0—6.6] | 0/129 | 0.0% [0.0—2.9] |
|  | 7F | 22/248 | 8.9% [5.9—13.1] | 49/536 | 9.1% [7.0—11.9] | 10/534 | 1.9% [1.0—3.4] | 0/54 | 0.0% [0.0—6.6] | 0/129 | 0.0% [0.0—2.9] |
|  | 19A | 20/248 | 8.1% [5.3—12.1] | 34/536 | 6.3% [4.6—8.7] | 29/534 | 5.4% [3.8—7.7] | 1/54 | 1.9% [0.3—9.8] | 7/129 | 5.4% [2.7—10.8] |
| PCV13 | total | 158/248 | 63.7% [57.6—69.4] | 195/536 | 36.4% [32.4—40.5] | 113/534 | 21.2% [17.9—24.8] | 6/54 | 11.1% [5.2—22.2] | 37/129 | 28.7% [21.6—37.0] |
| Non-PCV13 serotype | total | 90/248 | 36.3% [30.6—42.4] | 341/536 | 63.6% [59.5—67.6] | 421/534 | 78.8% [75.2—82.1] | 48/54 | 88.9% [77.8—94.8] | 92/129 | 71.3% [63.0—78.4] |
| PCV15-13 | subtotal | 29/248 | 11.7% [8.3—16.3] | 63/536 | 11.8% [9.3—14.8] | 62/534 | 11.6% [9.2—14.6] | 4/54 | 7.4% [2.9—17.6] | 8/129 | 6.2% [3.2—11.8] |
|  | 22F | 17/248 | 6.9% [4.3—10.7] | 44/536 | 8.2% [6.2—10.8] | 42/534 | 7.9% [5.9—10.5] | 4/54 | 7.4% [2.9—17.6] | 8/129 | 6.2% [3.2—11.8] |
|  | 33F | 12/248 | 4.8% [2.8—8.3] | 19/536 | 3.5% [2.3—5.5] | 20/534 | 3.7% [2.4—5.7] | 0/54 | 0.0% [0.0—6.6] | 0/129 | 0.0% [0.0—2.9] |
| PCV15 | total | 187/248 | 75.4% [69.7—80.3] | 258/536 | 48.1% [43.9—52.4] | 175/534 | 32.8% [28.9—36.9] | 10/54 | 18.5% [10.4—30.8] | 45/129 | 34.9% [27.2—43.4] |
| Non-PCV15 serotype | total | 61/248 | 24.6% [19.7—30.3] | 278/536 | 51.9% [47.6—56.1] | 359/534 | 67.2% [63.1—71.1] | 44/54 | 81.5% [69.2—89.6] | 84/129 | 65.1% [56.6—72.8] |
| PCV20-15 | subtotal | 38/248 | 15.3% [11.4—20.3] | 133/536 | 24.8% [21.3—28.6] | 221/534 | 41.4% [37.3—45.6] | 17/54 | 31.5% [20.7—44.7] | 43/129 | 33.3% [25.8—41.8] |
|  | 8 | 18/248 | 7.3% [4.6—11.2] | 62/536 | 11.6% [9.1—14.6] | 113/534 | 21.2% [17.9—24.8] | 9/54 | 16.7% [9.0—28.7] | 33/129 | 25.6% [18.8—33.7] |
|  | 10A | 0/248 | 0.0% [0.0—1.5] | 11/536 | 2.1% [1.1—3.6] | 19/534 | 3.6% [2.3—5.5] | 2/54 | 3.7% [1.0—12.5] | 2/129 | 1.6% [0.4—5.5] |
|  | 11A | 6/248 | 2.4% [1.1—5.2] | 7/536 | 1.3% [0.6—2.7] | 23/534 | 4.3% [2.9—6.4] | 2/54 | 3.7% [1.0—12.5] | 4/129 | 3.1% [1.2—7.7] |
|  | 12F | 10/248 | 4.0% [2.2—7.3] | 48/536 | 9.0% [6.8—11.7] | 57/534 | 10.7% [8.3—13.6] | 3/54 | 5.6% [1.9—15.1] | 3/129 | 2.3% [0.8—6.6] |
|  | 15B | 4/248 | 1.6% [0.6—4.1] | 5/536 | 0.9% [0.4—2.2] | 9/534 | 1.7% [0.9—3.2] | 1/54 | 1.9% [0.3—9.8] | 1/129 | 0.8% [0.1—4.3] |
| PCV20-13 | total | 67/248 | 27.0% [21.9—32.9] | 196/536 | 36.6% [32.6—40.7] | 283/534 | 53.0% [48.8—57.2] | 21/54 | 38.9% [27.0—52.2] | 51/129 | 39.5% [31.5—48.2] |
| PCV20 | total | 225/248 | 90.7% [86.5—93.7] | 391/536 | 72.9% [69.0—76.5] | 396/534 | 74.2% [70.3—77.7] | 27/54 | 50.0% [37.1—62.9] | 88/129 | 68.2% [59.8—75.6] |
| Non-PCV serotype | total | 23/248 | 9.3% [6.3—13.5] | 145/536 | 27.1% [23.5—31.0] | 138/534 | 25.8% [22.3—29.7] | 27/54 | 50.0% [37.1—62.9] | 41/129 | 31.8% [24.4—40.2] |
|  | 2 | 0/248 | 0.0% [0.0—1.5] | 0/536 | 0.0% [0.0—0.7] | 1/534 | 0.2% [0.0—1.1] | 0/54 | 0.0% [0.0—6.6] | 0/129 | 0.0% [0.0—2.9] |
|  | 9N | 5/248 | 2.0% [0.9—4.6] | 20/536 | 3.7% [2.4—5.7] | 27/534 | 5.1% [3.5—7.3] | 3/54 | 5.6% [1.9—15.1] | 10/129 | 7.8% [4.3—13.7] |
|  | 17F | 0/248 | 0.0% [0.0—1.5] | 0/536 | 0.0% [0.0—0.7] | 6/534 | 1.1% [0.5—2.4] | 0/54 | 0.0% [0.0—6.6] | 2/129 | 1.6% [0.4—5.5] |
|  | 20 | 1/248 | 0.4% [0.1—2.2] | 4/536 | 0.7% [0.3—1.9] | 9/534 | 1.7% [0.9—3.2] | 4/54 | 7.4% [2.9—17.6] | 4/129 | 3.1% [1.2—7.7] |
|  | 6C | 2/248 | 0.8% [0.2—2.9] | 23/536 | 4.3% [2.9—6.4] | 3/534 | 0.6% [0.2—1.6] | 1/54 | 1.9% [0.3—9.8] | 1/129 | 0.8% [0.1—4.3] |
|  | 7C | 0/248 | 0.0% [0.0—1.5] | 0/536 | 0.0% [0.0—0.7] | 5/534 | 0.9% [0.4—2.2] | 1/54 | 1.9% [0.3—9.8] | 5/129 | 3.9% [1.7—8.8] |

|  |  |  |  |  |  |  |  |  |  |  |
| --- | --- | --- | --- | --- | --- | --- | --- | --- | --- | --- |
| 10B | 0/248 | 0.0% [0.0—1.5] | 0/536 | 0.0% [0.0—0.7] | 2/534 | 0.4% [0.1—1.4] | 0/54 | 0.0% [0.0—6.6] | 0/129 | 0.0% [0.0—2.9] |
| 12B | 0/248 | 0.0% [0.0—1.5] | 3/536 | 0.6% [0.2—1.6] | 0/534 | 0.0% [0.0—0.7] | 0/54 | 0.0% [0.0—6.6] | 0/129 | 0.0% [0.0—2.9] |
| 13 | 0/248 | 0.0% [0.0—1.5] | 1/536 | 0.2% [0.0—1.0] | 1/534 | 0.2% [0.0—1.1] | 0/54 | 0.0% [0.0—6.6] | 0/129 | 0.0% [0.0—2.9] |
| 15A | 2/248 | 0.8% [0.2—2.9] | 24/536 | 4.5% [3.0—6.6] | 20/534 | 3.7% [2.4—5.7] | 2/54 | 3.7% [1.0—12.5] | 5/129 | 3.9% [1.7—8.8] |
| 15C | 3/248 | 1.2% [0.4—3.5] | 2/536 | 0.4% [0.1—1.4] | 1/534 | 0.2% [0.0—1.1] | 0/54 | 0.0% [0.0—6.6] | 0/129 | 0.0% [0.0—2.9] |
| 16F | 2/248 | 0.8% [0.2—2.9] | 8/536 | 1.5% [0.8—2.9] | 11/534 | 2.1% [1.2—3.7] | 4/54 | 7.4% [2.9—17.6] | 1/129 | 0.8% [0.1—4.3] |
| 18A | 0/248 | 0.0% [0.0—1.5] | 0/536 | 0.0% [0.0—0.7] | 1/534 | 0.2% [0.0—1.1] | 0/54 | 0.0% [0.0—6.6] | 0/129 | 0.0% [0.0—2.9] |
| 21 | 0/248 | 0.0% [0.0—1.5] | 1/536 | 0.2% [0.0—1.0] | 1/534 | 0.2% [0.0—1.1] | 0/54 | 0.0% [0.0—6.6] | 0/129 | 0.0% [0.0—2.9] |
| 22A | 1/248 | 0.4% [0.1—2.2] | 0/536 | 0.0% [0.0—0.7] | 1/534 | 0.2% [0.0—1.1] | 0/54 | 0.0% [0.0—6.6] | 0/129 | 0.0% [0.0—2.9] |
| 23A | 1/248 | 0.4% [0.1—2.2] | 14/536 | 2.6% [1.6—4.3] | 8/534 | 1.5% [0.8—2.9] | 2/54 | 3.7% [1.0—12.5] | 6/129 | 4.7% [2.1—9.8] |
| 23B | 0/248 | 0.0% [0.0—1.5] | 11/536 | 2.1% [1.1—3.6] | 6/534 | 1.1% [0.5—2.4] | 3/54 | 5.6% [1.9—15.1] | 2/129 | 1.6% [0.4—5.5] |
| 24 | 0/248 | 0.0% [0.0—1.5] | 0/536 | 0.0% [0.0—0.7] | 1/534 | 0.2% [0.0—1.1] | 0/54 | 0.0% [0.0—6.6] | 0/129 | 0.0% [0.0—2.9] |
| 24F | 0/248 | 0.0% [0.0—1.5] | 6/536 | 1.1% [0.5—2.4] | 9/534 | 1.7% [0.9—3.2] | 1/54 | 1.9% [0.3—9.8] | 1/129 | 0.8% [0.1—4.3] |
| 27 | 0/248 | 0.0% [0.0—1.5] | 0/536 | 0.0% [0.0—0.7] | 2/534 | 0.4% [0.1—1.4] | 0/54 | 0.0% [0.0—6.6] | 1/129 | 0.8% [0.1—4.3] |
| 28A | 0/248 | 0.0% [0.0—1.5] | 0/536 | 0.0% [0.0—0.7] | 0/534 | 0.0% [0.0—0.7] | 1/54 | 1.9% [0.3—9.8] | 0/129 | 0.0% [0.0—2.9] |
| 31 | 3/248 | 1.2% [0.4—3.5] | 1/536 | 0.2% [0.0—1.0] | 2/534 | 0.4% [0.1—1.4] | 1/54 | 1.9% [0.3—9.8] | 0/129 | 0.0% [0.0—2.9] |
| 32B | 0/248 | 0.0% [0.0—1.5] | 1/536 | 0.2% [0.0—1.0] | 1/534 | 0.2% [0.0—1.1] | 0/54 | 0.0% [0.0—6.6] | 0/129 | 0.0% [0.0—2.9] |
| 33D | 0/248 | 0.0% [0.0—1.5] | 0/536 | 0.0% [0.0—0.7] | 2/534 | 0.4% [0.1—1.4] | 0/54 | 0.0% [0.0—6.6] | 0/129 | 0.0% [0.0—2.9] |
| 34 | 0/248 | 0.0% [0.0—1.5] | 0/536 | 0.0% [0.0—0.7] | 2/534 | 0.4% [0.1—1.4] | 0/54 | 0.0% [0.0—6.6] | 0/129 | 0.0% [0.0—2.9] |
| 35B | 1/248 | 0.4% [0.1—2.2] | 8/536 | 1.5% [0.8—2.9] | 11/534 | 2.1% [1.2—3.7] | 1/54 | 1.9% [0.3—9.8] | 2/129 | 1.6% [0.4—5.5] |
| 35C | 0/248 | 0.0% [0.0—1.5] | 2/536 | 0.4% [0.1—1.4] | 0/534 | 0.0% [0.0—0.7] | 0/54 | 0.0% [0.0—6.6] | 0/129 | 0.0% [0.0—2.9] |
| 35F | 0/248 | 0.0% [0.0—1.5] | 10/536 | 1.9% [1.0—3.4] | 3/534 | 0.6% [0.2—1.6] | 2/54 | 3.7% [1.0—12.5] | 1/129 | 0.8% [0.1—4.3] |
| 37 | 0/248 | 0.0% [0.0—1.5] | 1/536 | 0.2% [0.0—1.0] | 0/534 | 0.0% [0.0—0.7] | 1/54 | 1.9% [0.3—9.8] | 0/129 | 0.0% [0.0—2.9] |
| 38 | 2/248 | 0.8% [0.2—2.9] | 5/536 | 0.9% [0.4—2.2] | 2/534 | 0.4% [0.1—1.4] | 0/54 | 0.0% [0.0—6.6] | 0/129 | 0.0% [0.0—2.9] |

The incidence of invasive pneumococcal disease among hospitalised adults showed a slight upward trend between 2006 and late 2019 (Figure 1A) . During the early part of the COVID-19 pandemic in 2020, pneumococcal disease incidence suddenly declined. Over 2022 it gradually increased so that by December 2022 it had returned to a level similar to that observed before the COVID-19 pandemic (Figure 1A). Non-invasive pneumococcal disease showed a broadly similar pattern but is harder to interpret due to changing BinaxNOW testing patterns over the study period (Figure 1C and Supplementary Figure 3 and 4). PCV7-serotype disease decreased from 29.4% [24.1–35.4] in 2006-2009 to 7.0% [3.7–12.7] of serotype-known disease in 2021-2022, with PCV7-serotypes causing minimal disease from mid-2017 until their re-emergence during the COVID-19 pandemic. PCV13-7 disease represented 34.3% [28.6–40.4] of serotype-known disease in 2006-2009, decreased slightly to 29.3% [25.6–33.3] by 2010-2015, and continued to decrease to 21.7% [15.5–29.6] in 2021-2022, with serotypes 1, 3 and 19A persisting. In contrast, the proportion of PCV20-13 and non-PCV serotype disease increased during this period (Figure 1B). While blood culture testing rates remained stable over time, there were changes in UAT testing rates. Notably, the decreased pneumococcal disease incidence observed during the COVID-19 pandemic occurred without an equivalent decrease in BinaxNOW and blood culture testing over the same time period (Figure 1C). Following SARS-CoV-2 emergence, the most commonly identified serotypes were 3, 8, 9N, 19F, 19A and 22F (Figure 2A,B). In 2022, 6.2% [3.2–11.8], 33.3% [25.8–41.8], and 31.8% [24.4–40.2] of disease was attributable to PCV15-13, PCV20-15 or non-PCV serotypes respectively, with a significant proportion of disease arising from serotypes in current PCVs (Table Two). Among known-serotype cases, PCV7 and PCV13-7 serotypes represented 7.0% [3.7–12.7] and 21.7% [15.5– 29.6] of pneumococcal disease (Table Two). Overall, the proportion of disease caused by 19F, 19A and 22F remained relatively stable throughout the study, while a notable expansion of serotypes 3 and 8 disease occurred, alongside a slight increase in serotype 9N (Figure 2C).

**Figure One:**
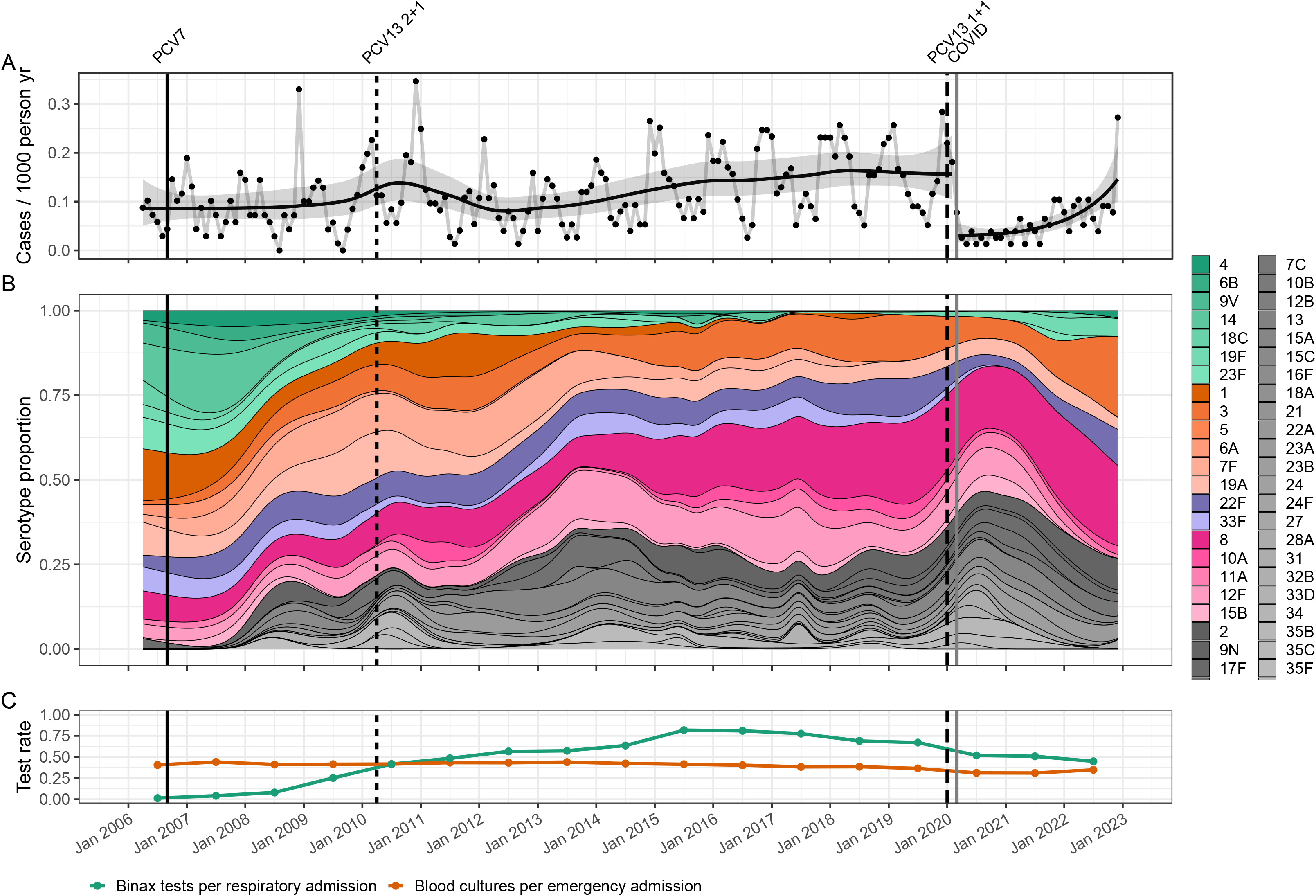
Distribution of pneumococcal serotypes in hospitalised patients. (A) Incidence of blood culture positive pneumococcal disease in Bristol, UK, 2006-2022. Data shown include only invasive disease (for comparison with non-invasive disease see Supplementary Data 4). Black dots represent monthly observations and the black line a binomial time series model with grey bars representing 95% confidence intervals. Population estimates are provided in Supplementary Data 4. (B) Multinomial proportion of pneumococcal serotypes, in the invasive pneumococcal disease population with known serotype. Green bars represent PCV7 serotypes; orange, PCV13-7; purple, PCV15-13; pink, PCV20-15; grey, serotypes not contained in PCV vaccines. Individual serotypes are shown in the legend on the panel.(C) The testing rates for blood cultures with respect to acute admissions and BinaxNOW tests with respect to acute respiratory admissions. Estimates are presented at the mid-year time point, relative to the data they represent. For all panels, vertical lines indicate the introduction of PCV7 (solid black), PCV13 (2+1 schedule) (thin black dash), PCV13 (1+1 schedule) (thick black dash) into the UK childhood vaccination programme and the beginning of significant hospital admissions in Bristol due to SARS-CoV-2 (grey solid).

**Figure Two:**
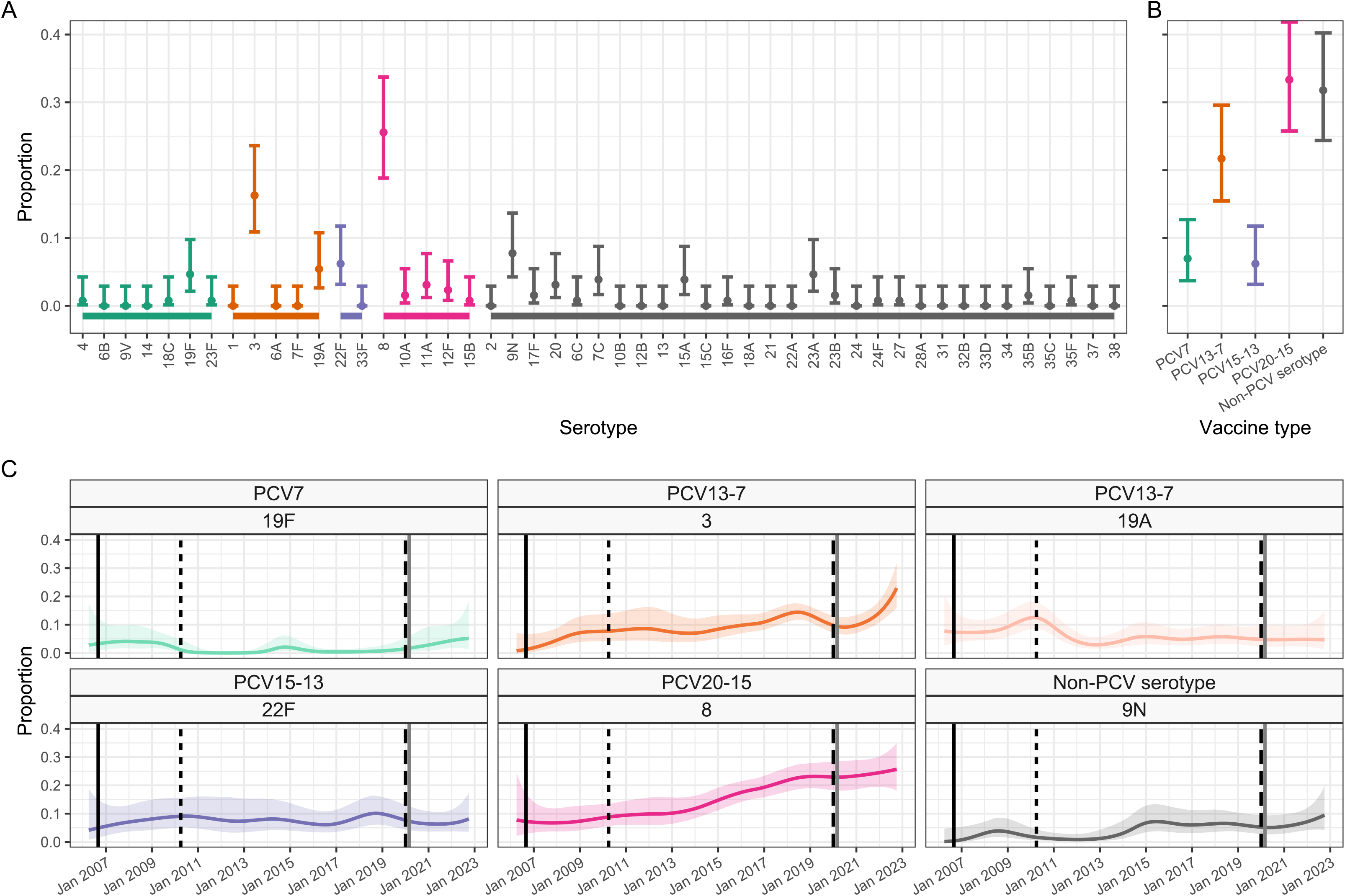
Distribution of pneumococcal serotypes in hospitalised patients following the emergence of SARS-CoV-2. The proportion of pneumococcal disease attributable to (A) each serotype and (B) by PCV vaccine group in Bristol, UK, from the 129 cases admitted between 1st Jan 2021 and 31st Dec 2022. Further data and time points are available in Table Two. (C) Binomial (one versus others) time series models showing the proportion of disease from Jan 2006 - Dec 2022 which was attributable to each of the six most common serotypes (3, 8, 9N, 19F, 19A, 22F). Solid lines represent smoothed point estimates with 95% confidence intervals shown as pale coloured areas. The confidence intervals in panels A&B do not coincide exactly with those in C as they represent different length time periods and use different models. For all panels, green bars represent PCV7 serotypes; orange, PCV13-7; purple, PCV15-13; pink, PCV20-15; grey, serotypes not contained in PCV vaccines).

Before the emergence of SARS-CoV-2, the age of patients admitted with invasive pneumococcal disease increased over time (Figure 3A) while disease severity decreased, as shown by the average CURB65 severity scores on admission (Figure 3B). ICU admission rates increased in earlier years and then plateaued, while inpatient mortality decreased and length of hospital admission remained broadly stable (Figure 3C-E). These trends were disrupted during the early stages of the SARS-CoV-2 pandemic and largely reverted to their previous trajectories in 2021-22. The proportion of patients aged 45-55y trended upwards, as compared to the pre-pandemic period, and the proportion of IPD cases requiring ICU admission also increased but these trends are based on small numbers. We could not account for the increase in ICU admissions seen following SARS-CoV-2 emergence through change in bed availability or clinical care pathways.

**Figure Three:**
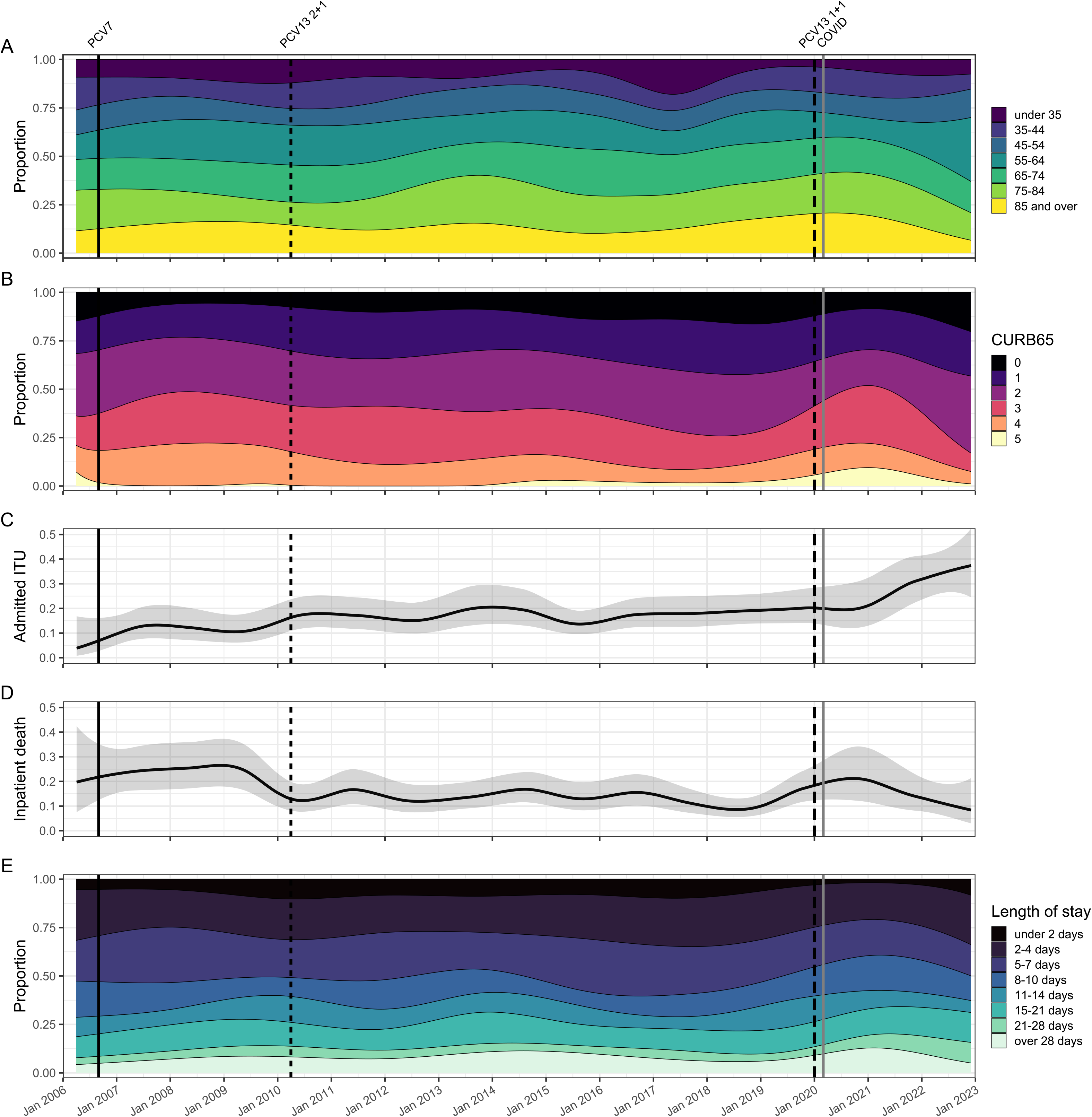
Severity of presentation and outcome of invasive pneumococcal disease over time. The proportion of invasive pneumococcal disease in hospitalised adults by (A) age category, (B) CURB65 score, (C) ICU admissions, (D) inpatient mortality and (E) hospital admission length. For panels (A), (B) and (E), categories are shown in the legend adjacent to each panel. The solid line in the multinomial time series models shown in panels (C) and (D) represent binomial time series models with 95% confidence intervals illustrated as grey area. Across all panels, the vertical lines denote the dates of PCV7 (solid black) and PCV13 2+1 (thin black dash) and 1+1 (thick dash black) schedule vaccine introduction and SARS-CoV-2 emergence (solid grey). Results for non-invasive pneumococcal disease are shown in Supplementary Data 4.

## DISCUSSION

This study examining hospitalised adults with pneumococcal disease in Bristol and Bath encompasses 17 years of PCV implementation into the UK childhood vaccination programme, and includes three years of the COVID-19 pandemic. While the proportion of IPD caused by PCV serotypes has declined across all age groups, a significant fraction of adult pneumococcal disease is still caused by PCV7 (7.0% [3.7–12.7]) and PCV13-7 (21.7% [15.5–29.6]) serotypes [5,23]. Additionally, the proportion of adult disease due to these serotypes has increased since COVID-19 pandemic restrictions began in March 2020, which also coincided with a change in the UK childhood immunisation schedule from three to two doses of PCV13 since April 2020. The persistence of PCV13 and PCV7-serotype disease in adults in this study occurs in the context of a long-standing, high coverage national childhood vaccination programme. This suggests that although giving PCVs to most children has had some indirect effects, these do not completely protect adults from PCV-serotype disease. The persistence of some PCV13 serotypes is concerning because they are associated with more severe disease and deaths in adults. Additionally, we have observed large increases in the proportion of non-PCV13 serotype disease since PCV13 implementation, consistent with national surveillance throughout the UK and Europe [5-7,12, 23]. In 2016-2019 and 2021-2022, 78.8% [75.2–82.1] and 71.3% [63.0–78.4] of cases among those with identified serotypes were due to non-PCV13 serotypes, respectively. In 2021-22, PCV20-13 serotypes accounted for 39.5% [31.5–48.2] of adult disease with identified serotypes.

Despite indirect effects operating to some degree, PCV serotypes continue to cause adult disease which is severe enough to require hospital admission. Although PCV7 serotypes remain rare causes of adult pneumococcal disease, serotype 19F has re-emerged. We find that most residual PCV serotype disease is due to PCV13-7 serotypes, especially serotypes 3 and 19A; observations which align with other reports [5,7,12,23]. The continued circulation of serotype 3, especially after the pandemic restrictions were lifted, is important, as this serotype is associated with more severe disease, especially in older adults. Ongoing circulation may, in part, be due to clade II expansion following clade distribution shift [24]. Serotype 19A is also of concern because it has previously been associated with high antibiotic resistance rates in some countries [25]. Indirect effects of PCVs are thought to occur via reduction in nasopharyngeal carriage and density of vaccine serotypes in vaccinated children, which interrupts transmission to both vaccinated and unvaccinated contacts [2–4]. Generally, sustained high PCV coverage in children is assumed to result in reliable indirect effects in adults: notably, the UK 1+1 PCV13 childhood immunisation schedule relies, to a degree, on these indirect effects, to protect infants who only receive one priming PCV dose before their one-year booster [26]. However, we cannot explain the recent proportional increase in PCV13-serotype disease we observed by falling paediatric PCV vaccination rates. In the UK, vaccination is offered free of charge and associated with high uptake and PCV uptake was >90% and >94% for the first and second PCV dose by 12-months and 24-months in South-West England in 2021-22, respectively [27].

Concerning, too, is the high proportion of cases that are currently not vaccine preventable: during 2016-19, PCV20-13 and non-PCV serotypes represented 53.0% [48.8–57.2] and 25.8% [22.3–29.7], changing to 39.5% [31.5–48.2] and 31.8% [24.4–40.2] in 2021-2022, respectively. In 2021-22, PCV15 serotypes represented 34.9% [27.2–43.4] of IPD, PCV20 represented 68.2% [59.8–75.6] and non-PCV serotypes represented 31.8% [24.4–40.2] of total disease. Interestingly, although serotype replacement has been observed in both Europe and serotype distribution change in the US [5–7,21,28, 29]: reports from the US describe consistently stable rates of non-PCV13 serotype IPD after childhood PCV13 introduction [30]. Serotype replacement may occur through expansion of non-PCV strains or capsular switching of previously vaccine-preventable strains, in which recombination at the capsular gene locus results in serotype change [31]. In concordance with others, we find serotype 8 disease increasing, along with 23A and 9N [5,7,12].

Early in the COVID-19 pandemic we observed a disproportionate decrease in pneumococcal disease incidence, without a concomitant decrease in BinaxNOW and blood culture testing rates. This may have been due to reduced circulation and transmission of many respiratory pathogens, including pneumococcus, due to social distancing policy implementation [13,14,16], breaking transmission chains and affecting host-pathogen-pathogen interactions, so that severe pneumococcal infection became less common. Preceding viral infection is a major risk factor for pneumonia, and the relationships between influenza and RSV and pneumococcus are well described [32,33]. Pneumococcus may also modulate host immune responses to SARS-CoV-2 [34]. Prospective surveillance demonstrates that Europe experienced significant and sustained reductions in invasive disease due to pneumococcus, *H. influenzae*, and *N. meningitidis* in early 2020 [35]. Our study confirms that reduced testing practices in hospitals do not explain such reductions. However, changes in healthcare utilisation, may explain the incidence decreases that occurred. For example, vulnerable patients had fewer admissions to secondary care during the pandemic [36]. An increased admission threshold or avoidance of secondary care would increase community-level management of cases and reduce cases of pneumococcal disease presenting, and consequently being diagnosed, in hospitals. Since fewer investigations are performed in community-level care, such cases would be missed by surveillance that is reliant on microbiological testing.

By combining microbiological results with clinical data, we are able to report on both disease epidemiology and severity over time. We found some evidence of more severe disease especially following SARS-CoV-2 emergence. The observed disruption to the broadly stable previous trends in disease severity during the early part of the SARS-CoV-2 pandemic may be a result of observation bias, a consequence of the pandemic, or both. While ICU admission rates increased for pneumococcal disease, we observed a reduction in admission CURB65 severity scores, consistent with the 2018 BTS national pneumonia audit which also reported decreased admission CURB65 scores [37]. This apparent discrepancy might be explained by changes in patients or their treatment: for example, older patients with more severe disease may be less likely to receive ICU care than younger patients due to clinical prognostic decision making. Other recent changes in treatment, such as increased availability of positive pressure support or earlier antibiotic administration, may also have changed patient courses and outcomes, including severity and mortality. Significant changes in pneumococcal infection severity have occurred since 2020, including an increased proportion of adults aged 45-55y who required hospital treatment. The recent increase in cases is probably related to relaxation of COVID-19 restrictions. Recent studies highlight the critical role of respiratory viruses (e.g. RSV, human metapneumovirus, influenza) in contributing to pneumococcal disease [17]. Large, out of season viral outbreaks which occurred following easing of COVID-19 restrictions [38] are therefore likely to have contributed to increasing pneumococcal disease reported here, which will need careful surveillance in future years. Independent of changes in clinical practice, serotype distribution changes may affect patient outcomes and healthcare usage as invasive potential varies by serotype.

This study has many strengths. We identified patients with pneumococcal infection in a population of approximately one million adults. We report disease over a 17-year period with epidemiological data supported by detailed clinical information at an individualised patient level. Data on admission numbers and testing practices allow us to interpret incidence considering these factors, which might otherwise bias the estimate obtained. Linkage with the UKHSA national reference laboratory allows us to report serotype data whenever available, strengthening this study considerably. Importantly, our study also reports pneumococcal disease confirmed by UAT: this is not detected by national surveillance and allows us to estimate pneumococcal disease incidence and burden more accurately. The study also has some limitations. The UAT does not identify serotype: we were thus unable to report serotype data in 60% of our cohort; however, patients with known and unknown serotype disease had broadly similar demographic, clinical characteristics and outcomes. UAT has a sensitivity of 65% and test results may remain positive for some time after pneumococcal disease has been treated [39]. Although the data are concordant with national UKHSA epidemiological and BTS pneumonia audit data [37], providing significant reassurance, this regional study may not be representative of other regions or populations. As a retrospective observational study, only information as documented in medical records could be included. Patient treatment decisions may have varied over time and by treating physicians, and systemic changes in patient treatment, healthcare provision or admission threshold and microbiological testing practices may have occurred. These factors may have affected the trends reported in this analysis. The serotype trends reported in 2020-22 are based on substantially lower case numbers than those seen before the emergence of SARS-CoV-2, and ongoing monitoring of trends will be important as disease incidence adjusts following the COVID-19 pandemic.

In conclusion, this study provides evidence that PCV13 serotypes continue to cause disease in adults, suggesting indirect effects of the childhood PCV programme may not lead to further reductions in the current residual burden of these serotypes in adults. Following the COVID-19 pandemic, serotype distribution changes have resulted in an increase in adult pneumococcal disease attributable to PCV serotypes, and a significant burden of current disease could be directly prevented by introducing PCV20 which has recently been licensed for adults. Ongoing surveillance of adult pneumococcal disease is vital to evaluate vaccine impact and monitor replacement disease. The extent of indirect effects of PCVs should continue to be carefully evaluated and considered in formulating future public health policy recommendations.

## Data Availability

The data used in this study are sensitive and cannot be made publicly available without breaching patient confidentiality rules. Therefore, individual participant data and a data dictionary is not available to other researchers.

## Author Biography

Dr Catherine Hyams undertook her PhD in pneumococcal infection at UCL and is now a PostDoctoral Research Fellow at the University of Bristol. She undertakes research on respiratory infection such as pneumococcus, RSV and COVID-19, and currently holds roles within the British Thoracic Society Pulmonary Infection Special Advisory Group and UKHSA COVID-19 vaccine effectiveness working group.

## Declarations

CH is Principal Investigator of the AvonCAP study which is an investigator-led University of Bristol study funded by Pfizer. AF is a member of the Joint Committee on Vaccination and Immunization (JCVI) and was, until December 2022, chair of the World Health Organization European Technical Advisory Group of Experts on Immunization (ETAGE) committee. In addition to receiving funding from Pfizer as Chief Investigator of this study, he leads another project investigating transmission of respiratory bacteria in families jointly funded by Pfizer and the Gates Foundation. The other authors have no relevant conflicts of interest to declare.

## Contributions

CH, RCh, OMW, SL and AF generated the research questions and analysis plan. CH, RH, DH, GF, RCo, PN, PW, ZAC collected data. CH, RH, and DH verified the data. CH, RCh, LD, and AF undertook the data analysis. AF, OMW and AM provided oversight of the research. All authors contributed to the preparation of the manuscript and its revision for publication and had responsibility for the decision to publish.

## ACKNOWLEDGEMENTS

The authors would like to acknowledge the research teams at The Royal United, North Bristol and University Hospitals of Bristol and Weston NHS Trusts.

