## Supplementary Data for "Trends in serotype distribution and disease severity in adults hospitalised with *Streptococcus pneumoniae* infection in Bristol and Bath: a retrospective cohort study, 2006-2022"

**Supplementary Data 1: Serotypes contained within each pneumococcal vaccination and vaccine group.**

| **Vaccination** | **Serotypes** |
| --- | --- |
| PPV-23, PneumoVax® | 1, 2, 3, 4, 5, 6B, 7F, 8, 9N, 9V, 10A, 11A, 12F, 14, 15B, 17F, 18C, 19F, 19A, 20, 22F, 23F, 33F. |
| PCV-7, Prevenar® | 4, 6B, 9V, 14, 18C, 19F and 23F |
| PCV-13, Prevenar13® | 1, 3, 4, 5, 6A, 6B, 7F, 9V, 14, 18C, 19A, 19F, & 23F |
| PCV-15, VAXNEUVANCE® | 1, 3, 4, 5, 6A, 6B, 7F, 9V, 14, 18C, 19A, 19F, 22F, 23F and 33F |
| PCV-20, Prevenar20® | 1, 3, 4, 5, 6A, 6B, 7F, 8, 9V,10A, 11A, 12F, 14, 15B, 18C, 19A, 19F, 22F, 23F and 33F |
| PCV13-7 serotypes | 1, 3, 5, 6A, 7F, 19A |
| PCV15-13 serotypes | 22F, 33F |
| PCV20-15 serotypes | 8, 10A, 11A, 12F, 15B |
| PCV20-13 serotypes | 8, 10A, 11A, 12F, 15B, 22F, 33F |
| Non-PCV serotypes | Any serotype not contained in PCV20 (and therefore PCV15, PCV13 and PCV7) |

PCV, Pneumococcal conjugate vaccine; PPV-23, Pneumococcal polysaccharide vaccine, 23 valent

**Supplementary Data 2: Demographic comparison between invasive and non-invasive pneumococcal disease**

|  |  | Invasive disease | Non-invasive disease |  |
| --- | --- | --- | --- | --- |
| Variable | Characteristic | Value (N=1686) | Value (N=2033) | P value |
| Age | Median [IQR] | 65.9 [50.8—79] | 66.3 [50—78.8] | 0.74 |
| Gender | Male % (N) | 50.2% (847) | 48.8% (993) | 0.41 |
|  | Female % (N) | 49.8% (839) | 51.2% (1040) |  |
| Serotype Status | Serotype identified % (N) | 89.0% (1501) | 0.0% (0) | <0.001 |
|  | No serotype % (N) | 11.0% (185) | 100.0% (2033) |  |
| Smoker | Non-smoker % (N) | 29.4% (495) | 29.4% (597) | 0.95 |
|  | Ex-smoker % (N) | 40.3% (680) | 40.8% (829) |  |
|  | Current smoker % (N) | 30.3% (511) | 29.9% (607) |  |
| CCI | Median [IQR] | 4 [1—6] | 4 [1—6] | 0.31 |
| Test Type | Blood culture only % (N) | 78.6% (1325) | 0.0% (0) | <0.001 |
|  | UAT only % (N) | 3.0% (51) | 100.0% (2033) |  |
|  | Blood culture and UAT % (N) | 17.5% (295) | 0.0% (0) |  |
|  | CSF PCR % (N) | 0.5% (8) | 0.0% (0) |  |
|  | Blood PCR % (N) | 0.4% (7) | 0.0% (0) |  |
| Infection Site | Lung % (N) | 84.2% (1419) | 99.2% (2017) | <0.001 |
|  | Meningitis % (N) | 10.2% (172) | 0.0% (0) |  |
|  | Septic arthritis % (N) | 2.1% (36) | 0.0% (0) |  |
|  | ENT % (N) | 0.9% (15) | 0.1% (2) |  |
|  | Other % (N) | 2.6% (44) | 0.7% (14) |  |
| PPV23 Vaccination | No % (N) | 57.4% (967) | 57.6% (1171) | 0.99 |
|  | Under 6 months % (N) | 3.6% (60) | 3.5% (72) |  |
|  | Over 6 months % (N) | 39.1% (659) | 38.8% (789) |  |
|  | <missing> % (N) | 0.0% (0) | 0.0% (1) |  |
| Non-Invasive Ventilation | yes % (N) | 2.3% (39) | 5.1% (103) | <0.001 |
| Intubation | yes % (N) | 14.0% (236) | 11.5% (234) | 0.026 |
| Inpatient Death | yes % (N) | 15.1% (255) | 12.4% (253) | 0.019 |
| Significance determined using Fisher's exact test (categorical variables) or 2 sample Kolmogorov-Smirnov test, 2 sample Wilcoxon Rank Sum test (continuous variables) Normality of distributions determined using Anderson-Darling normality test  CCI, Charlson comorbidity Index; CSF, cerebrospinal fluid; ENT, ear, nose and throat; IQR, interquartile range; PCR, polymerase chain reaction; PPV23, pneumococcal polysaccharide vaccine 23-valent; UAT, urinary antigen test | | | | |

**Supplementary Data 3: Test positive pneumococcal cases in both invasive and non-invasive cohorts by test method**

The incidence of detected pneumococcal disease is mostly stable for blood cultures, and blood culture testing rate has been stable over time, but “other” testing (largely BinaxNOW) has increased over the study period (see Figure 1C in the main paper) and hence there is an ascertainment bias in the case numbers in the non-invasive (i.e. Test type: Other) cohort.

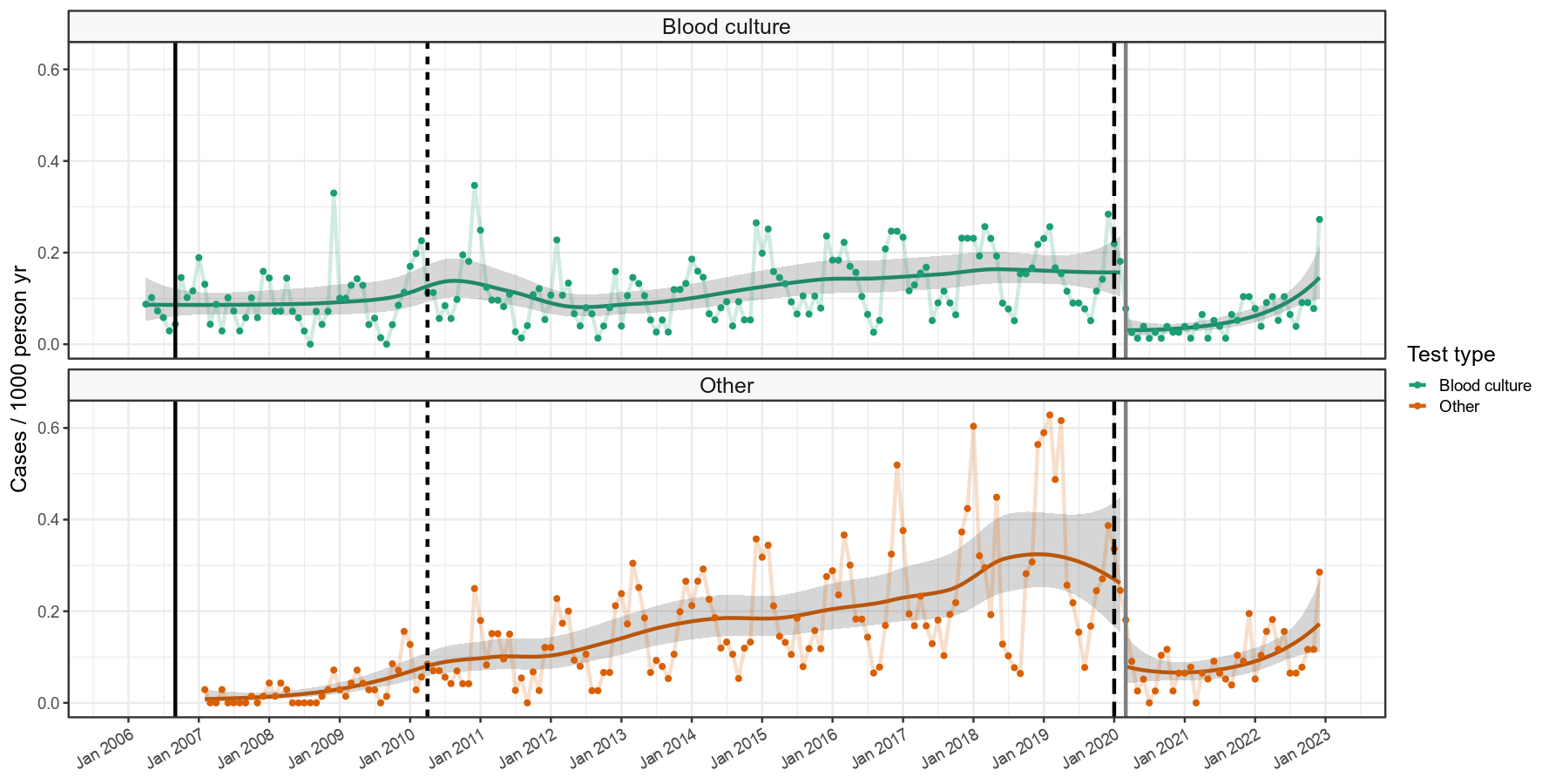

**Supplementary Data 4: Test positive pneumococcal cases in invasive and non-invasive cohorts by test method and CURB65 score**

Ascertainment bias in the principally non-invasive (i.e. Test type: Other) cohort is not clearly related to severity, and proportionality through time is broadly similar for different CURB65 scores. Severity analysis in the main paper is focussed only on invasive disease and hence is not affected by these changes in ascertainment over the study period.

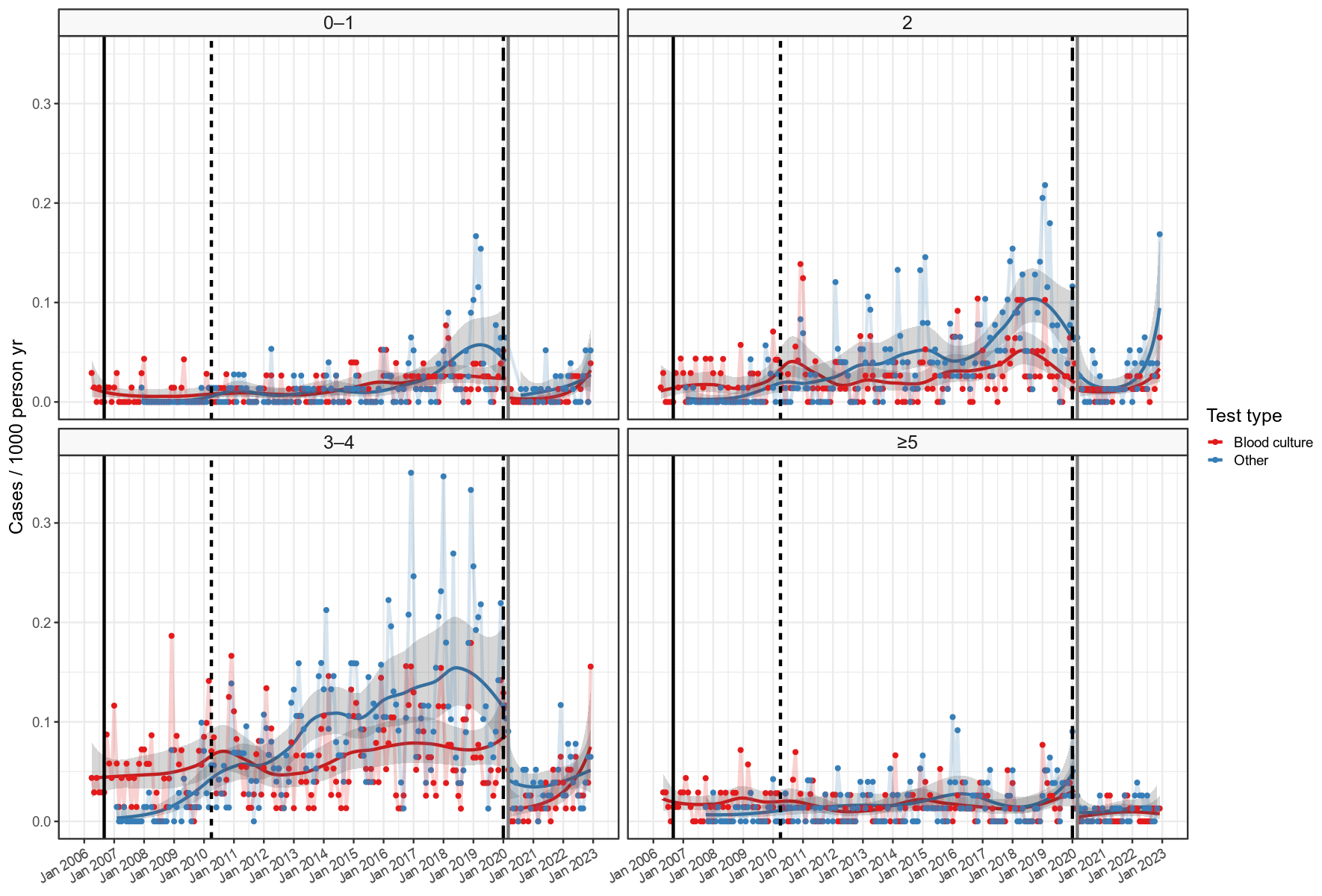

**Supplementary Data 5: Population estimates in the over 16 age groups**

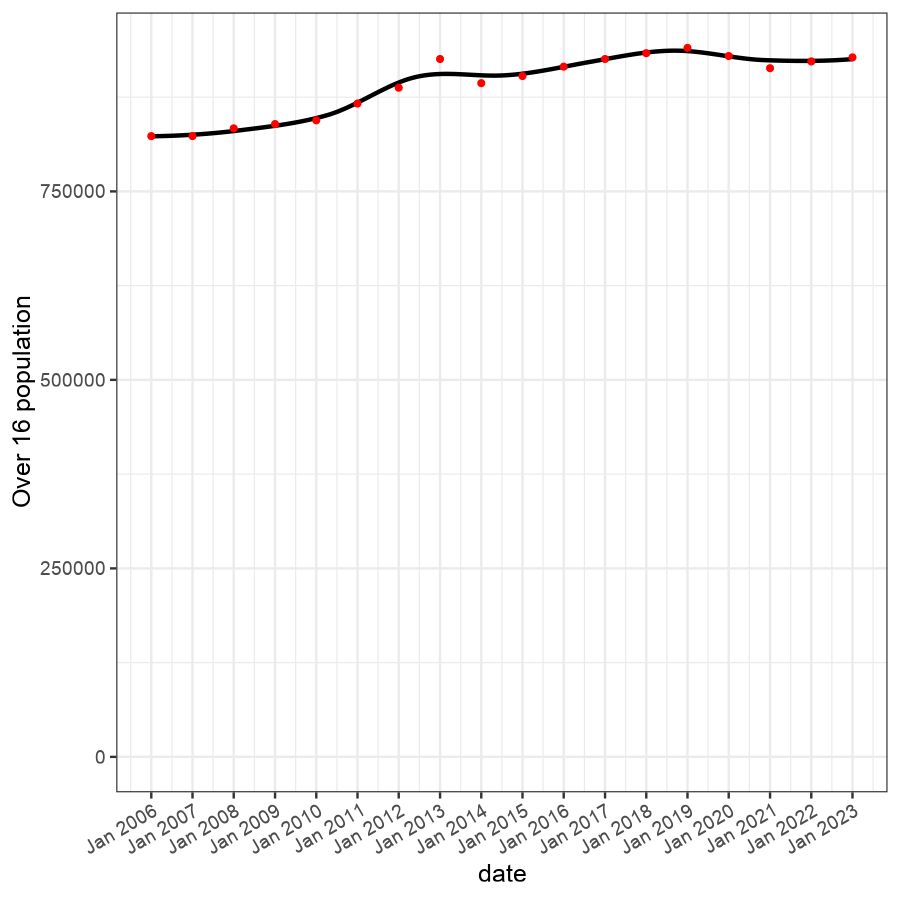

**Supplementary Data 6: Severity of presentation and outcome of non-invasive pneumococcal disease over time**

The proportion of non-invasive pneumococcal disease in hospitalised adults by (A) age category, (B) CURB65 score, (C) ICU admissions, (D) inpatient mortality and (E) hospital admission length. For panels (A), (B) and (E), categories are provided in the legend adjacent to each panel. The solid line in the multinomial time series models shown in panels (C) and (D) represent binomial time series models with 95% confidence intervals illustrated as grey area. Across all panels, the dates of PCV7 and PCV13 (2+1 and 1+1 schedule) vaccine introduction and SARS-CoV-2 emergence are shown. As this figure relies on UAT data the early part of the time series should be interpreted with caution as routine use was not fully established until around 2010 and significant ascertainment bias exists.

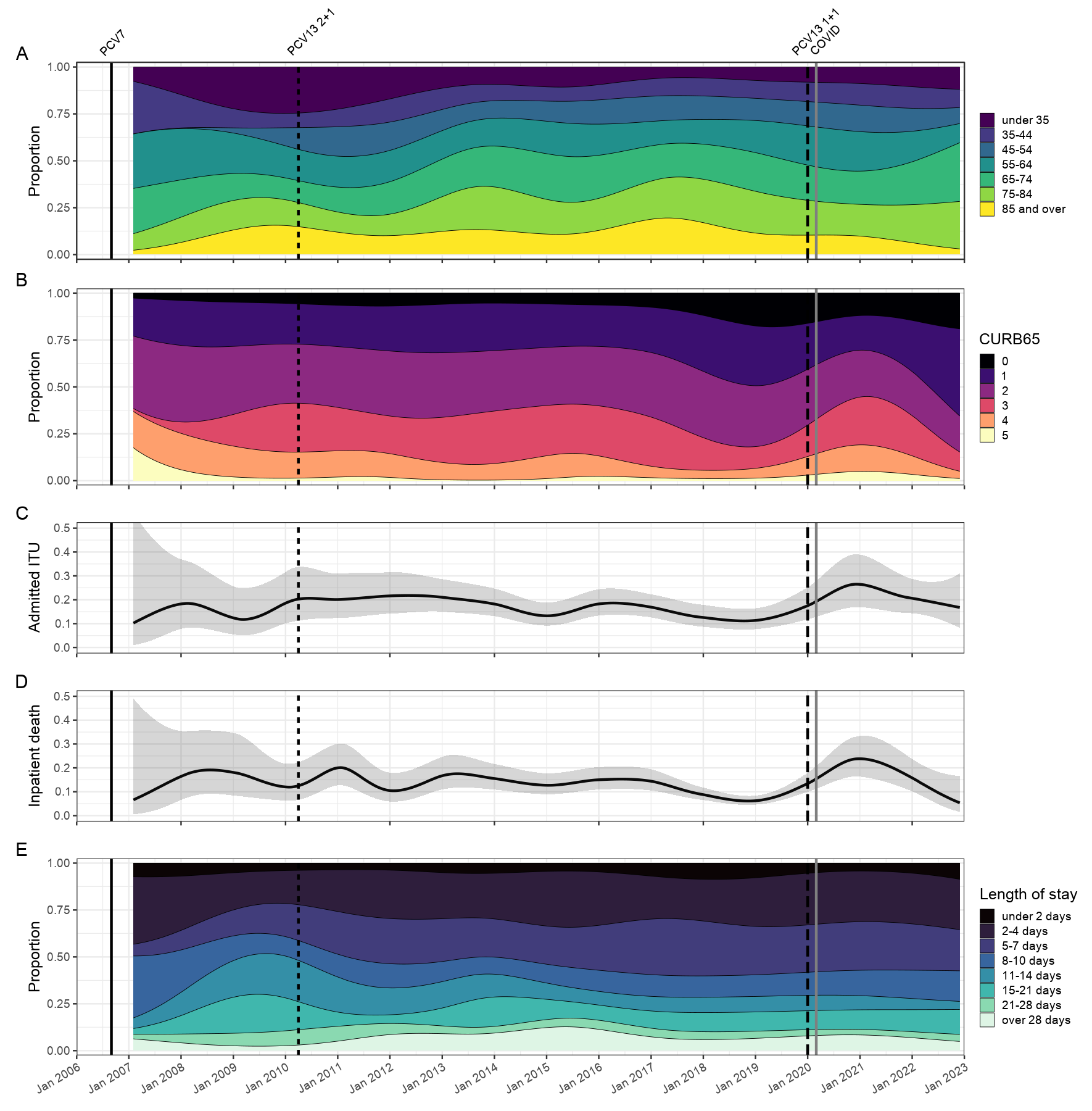
